# A qualitative study exploring how secondary school PE uniform policies influence body image, and PE engagement among adolescent girls

**DOI:** 10.1101/2024.12.19.24319312

**Authors:** Alice Porter, Elin Cawley, Laura Chapman, Charlotte Crisp, Ruth Wadman, Sally Barber, Ian Penton-Voak, Angela Attwood, Russell Jago, Helen Bould

**Author notes:** **Correspondence to**: Alice Porter, Population Health Sciences, Bristol Medical School, Canynge Hall, 39 Whatley Road, Bristol, BS8 2PS, UK.

## Abstract

**Background:** Many adolescent girls experience body dissatisfaction and have low levels of physical activity. Secondary school Physical Education (PE) offers opportunities for girls to build self-confidence and stay active; however, PE uniforms can be a barrier to participation.

**Objectives:** To explore how secondary school PE uniform policies influence body image and PE engagement (participation and enjoyment) among adolescent girls, and how these policies could be co-developed in future.

**Design:** A qualitative study involving focus groups and interviews.

**Participants and setting:** Forty-four 13-14-year-old girls and six PE staff members from six mixed-sex secondary schools in England.

**Data collection and analysis:** Using topic guides and participatory activities to aid discussions, we explored PE uniform preferences and the influence on body image and PE engagement with adolescent girls, as well as the PE uniform policy development process with PE staff. Data were analysed using reflexive thematic analysis, supported by NVivo 14.

**Results:** Three themes were generated. Theme 1, ‘Striking the right balance between choice, comfort and uniformity’, describes the challenges of developing PE uniform policies that offer pupils choice to maximise comfort, whilst maintaining uniformity to ensure smartness, and to reduce social comparison. Theme 2, ‘PE uniforms are “made for boys”’ reflects that current policies can often provide unisex uniforms that don’t fit the female body, or gendered options that limit girls’ choices over style and fit. Theme 3, ‘Self-confidence influences comfort in wearing PE uniform, and in turn PE engagement’ suggests girls with high self-confidence may be less concerned about others’ opinions and how they look, leading to greater PE enjoyment, whereas girls with lower self-confidence described feeling self-conscious, especially in communal changing rooms, which could impact their comfort and PE engagement.

**Conclusions:** Our findings suggest that developing PE uniform policies, which allow pupils to choose their own bottoms, wear additional layers, and wear PE uniform all day may improve comfort and inclusivity among girls, facilitating better PE engagement.

**Strengths and limitations of this study:**

- Our qualitative approach, using participatory activities to prompt discussions, enabled in-depth exploration into how PE uniforms can influence body image and PE engagement among adolescent girls.
- Data from adolescent girls and PE staff were triangulated to ensure PE uniform policy recommendations considered both pupil and staff perspectives.
- Due to challenges with recruiting schools, our sample was made up of mixed-sex, mainly affluent schools in South West England only.
- We had limited representation from pupils of ethnic minority backgrounds and those with low levels of PE enjoyment.

## INTRODUCTION

Adolescence is a critical period of vulnerability for the development of mental health problems.^1^ ^2^ Physical and hormonal changes during adolescence, coupled with heightened social pressures and increased social media use, can contribute to poor body image and low self-esteem.^3^ ^4^ Body image is defined as “the perception that individuals have of their physical self and the thoughts and feelings that result from that perception”.^5^ Body dissatisfaction, characterized by negative thoughts and feelings regarding one’s body, (often stemming from a perceived discrepancy between one’s own body and an ’ideal’ body),^6^ is associated with increased risks of anxiety, depression, and the development of eating disorders, and is particularly prominent among adolescent girls.^7–10^

Physical activity has multiple benefits, including improved mental health, well-being, and self-esteem.^11^ ^12^ Despite these benefits, adolescence also often coincides with a reduction in physical activity.^13–15^ Gender inequality in physical activity emerges as early as two years, with research suggesting girls engage in less outdoor play than boys.^16^ However, this gap widens with age, as the decline in physical activity starts sooner, and becomes steeper for girls than boys from childhood into adolescence.^13^ ^14^ ^17–19^ Research indicates that girls can experience multiple barriers to continuing to engage in physical activity, including low self-confidence, poor body image, gender stereotypes and norms, safety concerns, insufficient peer and family support, and discomfort with available clothing options.^20^ ^21^ It is therefore important to enable and promote the inclusivity of physical activity and sport for girls.

As part of the national curriculum for education, secondary schools offer a space for adolescent girls to engage in physical activity through Physical Education (PE), which can be an opportunity to build competency in physical and social skills, increase self-confidence, and establish long-term participation in physical activity.^22^ However, research suggests many adolescent girls dislike PE due to the focus on competitive sport and feeling self-conscious in front of peers.^23^ In addition, questionnaires show that many girls dislike and feel uncomfortable in their PE uniform.^24–27^ For example, the 2024 Youth Sports Trust Active Girls survey demonstrated that only 21% of adolescent girls in England were satisfied with their PE uniform, and only 47% felt comfortable wearing it.^24^ A retrospective study that asked women to reflect on their experiences of PE in school suggested that PE uniforms were a significant barrier to physical activity and sport engagement and, for some, negatively impacted body image.^28^ However, there is a lack of in-depth qualitative research exploring how PE uniforms may influence body image and PE engagement among adolescent girls.

Government guidelines in England state that schools should “choose a PE kit which is practical, comfortable, appropriate to the activity involved and affordable”.^29^ Similar guidelines are provided in Scotland, Wales, Australia and New Zealand.^30–33^ These guidelines advocate for gender equality within school uniform policies, however details about how this can be achieved, as well as specific recommendations for PE uniform items are not provided. There is a need to develop more practical, evidence-based recommendations for school PE uniform policies to help schools improve the inclusivity of their PE uniforms. Therefore, the aims of this study were to:

- Explore whether and how secondary school PE uniform policies influence body image and PE engagement (i.e. participation and enjoyment) among adolescent girls.
- Explore the feasibility and acceptability of co-developing new PE uniform policies with school staff and pupils.

## METHODS

We conducted a qualitative study across six mixed-sex secondary schools in Bristol and South Gloucestershire, UK, via focus groups with Year 8 pupils and interviews with PE staff. Ethics approval was obtained from the Faculty of Health Sciences Ethics Committee at the University of Bristol, in February 2024 (Reference 17624) and informed written parental consent was provided for all children and informed consent for all PE staff.^34^ The study protocol was pre-registered on the Open Science Framework (OSF) (https://osf.io/krhg7). We followed the Standards for Reporting Qualitative Research (SRQR) (Supplementary Table 1).^35^

### Patient and public involvement and engagement (PPIE)

We conducted PPIE with adolescent girls, their parents, and secondary school stakeholders (teacher and Health and Education lead) to inform the development of topic guides and study documents. Following PPIE feedback, some questions were removed from the pupil topic guide due to being difficult to understand or not age-appropriate and suggestions were made to add interactive elements (see Study Procedures below). Additionally, study documents were refined to improve clarity.

### Participants and recruitment

We aimed to recruit 6-8 schools selected using publicly available UK government data.^36^ Recruitment focused on achieving diversity in school type (mixed-and single-sex, local authority and academy), school performance (based on Ofsted rating; single-word judgement given to schools following inspection, and Attainment 8 score; measure of average academic performance across eight subjects), and pupil demographics (proportion of children from ethnic minority backgrounds, and free school meal eligibility (an indicator of deprivation)). Private schools were excluded.

The inclusion criteria for focus groups were Year 8 pupils who identified as female or non-binary. We aimed to recruit pupils with varying PE enjoyment levels and from a range of ethnic backgrounds. PE staff with knowledge of the school PE uniform policy were included for interviews. Initial recruitment purposively targeted schools in Bristol/South Gloucestershire and Bradford, UK, based on the characteristics described above, however due to low response rates (only one targeted school responded), recruitment was expanded to include all secondary schools in our recruitment areas.

### Study Procedures

A recruitment email and information sheet were sent to all schools. Staff expressing interest received a participant information sheet and were asked to disseminate study materials (study advert, participant information sheet and consent form) to eligible pupils and parents.

Parental consent was obtained via an online form, which also collected demographic data on ethnicity, religion and family affluence (subjective proxy measure of income), and pupil-rated PE enjoyment level (one being low, five being high). To ensure all participants could contribute towards the discussion, we recruited a maximum of eight pupils per school. If more than eight pupils expressed interest, participants were purposively sampled based on PE enjoyment and ethnicity.

Six semi-structured face-to-face focus groups with pupils (without staff present) and five interviews with PE staff were conducted in schools between May and July 2024, by two female researchers (EC and AP or LC). One staff interview was conducted online via Teams by EC. Focus groups lasted between 33-49 minutes while interviews lasted between 35-45 minutes, and were audio-recorded using an encrypted device with participant consent. Pupils received a £20 gift voucher and staff received a £25 voucher as reimbursement for their time.

Topic guides (available in the protocol: https://osf.io/krhg7) were used for focus groups and interviews. School PE uniform policies, obtained via school websites prior to data collection were used to aid discussion, and are summarised in Supplementary Table 2. Focus group discussions covered pupils’ opinions on current PE uniform policies, their influence on PE engagement and body image, and interest in changing PE uniforms. Two activities were used to facilitate discussions: writing likes/dislikes about the PE uniform on Post-it notes and attaching them to their PE uniform items, which they had brought to the session; and indicating their level of self-confidence wearing their PE uniform by sticking a t-shirt (made from paper) onto a scale ranging from self-conscious to self-confident. PE staff interviews covered views on current PE uniform policies, the policy development process, influence on pupil PE engagement and body image, and feasibility and acceptability of co-developing new polices with pupils.

### Data analysis

Audio recordings were transcribed verbatim using a University-approved transcription service, and anonymised. Data included focus group and interview transcripts, and Post-it notes from pupil activities. We employed a reflexive thematic analysis (RTA)^37^ to interpret the data, supported by NVivo 14.^38^

The analysis process involved familiarisation with the data by anonymising, reading, and re-reading transcripts. This was followed by inductive coding to identify semantic (i.e. explicit, surface-level) and latent (i.e. underlying, interpretative) patterns of shared meaning across the data. To promote reflexivity, four researchers (AP, EC, LC, CC) independently coded four transcripts on two separate occasions (eight in total), who met to discuss their codes and interpretations of the data. The remaining transcripts were coded twice by AP and EC. Coding was iterative to support the theme generation process. Initial themes were generated using the codes, then reviewed and developed. These themes were refined through regular discussions between AP and EC and subsequently reviewed and discussed with the wider research team for further refinement. Theme descriptions were produced, providing quotes from transcripts to support the themes. In addition to RTA, the data was used to outline initial recommendations and key considerations for improving PE uniform policies for adolescent girls.

### Positionality

All researchers involved in coding the data were White females, highly educated to Masters or PhD level, with an interest in female health. AP, EC and LC had previous training and experience in qualitative analysis, and CC was new to qualitative analysis.

## RESULTS

### Sample

Six mixed-sex Academy schools in Bristol and South Gloucestershire were recruited, the majority of which were affluent schools. Table 1 presents the school characteristics. We had intended to recruit schools in Bradford, single-sex and local authority schools, as planned in our protocol, however were unable to due to lack of engagement from schools.

**Table 1.** Academy* school characteristics (N=6)

| Characteristic | Mean | Range |
| --- | --- | --- |
| Ofsted rating** | 2 | 1 - 3 |
| Percentage of pupils eligible for free school meals*** | 22 | 6.7 - 58 |
| Percentage of pupils classified as White British ethnicity | 66.5 | 42.4 - 82.2 |
| Attainment 8 score**** | 48 | 29.3 - 60.8 |

\*State-funded school that is independent from local authority
\*\*The Office for Standards in Education, Children's Services and Skills (Ofsted) ratings are a single-word judgement given to each education provider receiving a graded school inspection. These ratings help hold schools accountable for the education and care they provide to children. Ofsted ratings are represented as 1 - Outstanding, 2 - Good, 3 - Requires Improvement, 4 – Unsatisfactory.
\*\*\*Eligibility for Free School Meals is an indicator of deprivation (higher percentage representing greater deprivation). Average is 24.6% in England.<sup>36</sup>
\*\*\*\*A measure of pupil's average academic performance across eight subjects. Average is 48.6 for girls in England.<sup>39</sup>

Across the six schools, 58 Year 8 pupils (all female) expressed interest and 44 took part in focus groups (6-8 per school). Table 2 presents pupil characteristics, with the majority being of White ethnicity. Six PE staff (3 PE teachers, 3 Heads of PE) took part in interviews (one per school), four were female and two were male. Staff had secondary education experience ranging from seven to 29 years. A summary of the school PE uniform policies retrieved from school websites is presented in Supplementary Table 2, including discrepancies with how PE uniform policies were implemented in practice (i.e. comparing to focus group and interview data).

**Table 2.** Characteristics of participating Year 8 pupils (N=44)

| Characteristic | n | % |
| --- | --- | --- |
| Ethnicity* |  |  |
| White or White British | 29 | 66 |
| Mixed or multiple ethnic group | 2 | 5 |
| Arab | 1 | 2 |
| Not reported | 12 | 27 |
| Religion |  |  |
| Christian (Catholic, Protestant or any other Christian) | 16 | 36 |
| Muslim | 2 | 5 |
| No religion | 21 | 48 |
| Not reported | 5 | 11 |
| PE enjoyment level (1 being low and 5 being high) |  |  |
| 1 | 1 | 2 |
| 2 | 4 | 9 |
| 3 | 10 | 23 |
| 4 | 12 | 27 |
| 5 | 17 | 39 |
| Family affluence** |  |  |
| 1 | 0 | 0 |
| 2 | 3 | 7 |
| 3 | 23 | 52 |
| 4 | 11 | 25 |
| 5 | 0 | 0 |

\*Ethnicity was self-reported and then categorised using the UK government consensus categories.<sup>40</sup>
\*\*Parents were asked 'Compared to other families in the UK, would you say your family income is: 1 – Far below average (much lower than most other families), 2 – below average (lower than most other families), 3 – average (about the same as most other families), 4 – above average (more than most other families), 5 – far above average (much more than most other families).<sup>41</sup>

### Themes

Three themes were generated relating to how PE uniforms influence body image and PE engagement (enjoyment and participation) among female adolescents. These were 1) Striking the right balance between choice, comfort and uniformity, 2) PE uniforms are “made for boys”, and 3) Self-confidence influences comfort in wearing PE uniform, and in turn PE engagement.

### Theme 1: Striking the right balance between choice, comfort and uniformity

Findings highlighted the challenge of providing PE uniform policies that allow pupils choice in what they wear during PE lessons to maximise their comfort, whilst also maintaining a certain level of uniformity which ensures: 1) pupils look smart when representing their school; 2) the PE uniform can be worn by pupils of different gender identities (see link to Theme 2); and 3) girls look similar to their peers and do not stand out, which could exacerbate negative body image (see link to Theme 3).

> *“It’s what’s functional but also what looks smart, because they have to be comfortable in it.” (Female PE staff, School 4)*

Feeling comfortable in the PE uniform was important to both Year 8 girls and PE staff. Comfort was alluded to in two ways: 1) the physical comfort of the material and fit (e.g. thickness and type of material, not being restrictive) *(“It’s kind of like quite thick [the PE top material], so I normally wear like a thinner top and shorter type shorts because it’s just more comfortable.” Pupil, School 6*); and 2) psychological comfort *(“Because people might feel better in something slim fit than they do baggy because they like feel confident about their self and their body” Pupil, School 1).* It appeared that comfort was strongly influenced by level of choice, with a preference among the girls to *“be able to wear whatever we feel comfortable in” (Pupil, School 2).* Allowing choice, particularly over the style, fit and length of bottoms (e.g. leggings, shorts, joggers), and to wear additional layers (e.g. jackets, coats, underlayers) seemed to facilitate greater comfort.

> *“‘cause we have such a good like amount of choice, I think it’s easier to wear something you’re more comfortable in one day if you don’t really wanna wear something like that.” (Pupil, School 4)*

Looking good and being “on trend” also appeared to matter to some pupils, alongside comfort. Many girls wanted the option to wear cycle shorts, although some PE staff felt these choices were influenced more by fashion and social media, than comfort and practicality. Whilst pupils valued having options for better comfort, some seemed to worry about standing out and being picked on for it, especially when choosing the type of bottoms to wear and choosing colours (e.g. for trainers), as well as when having to wear spare uniform (if uniform was forgotten). It was noted by some pupils and PE staff that trends were set by the *“popular”* or *“sporty”* girls, which other girls may feel pressure to follow, even if they are less comfortable options (see links to Theme 3).

> *“if you decide to wear something that not everyone wears but it’s still within uniform restriction, a lot of people tend to say something about it and then even if that thing is meant to be positive or meant to be a joke, it still can make people feel insecure.” (Pupil, School 2)*

> *“say there was a skort, I feel like some people would wear the skort and then I’d feel bad for people who wear shorts because just knowing school in general. I feel like the popular ones would wear like skorts or something and then people would find it weird if you wore shorts” (Pupil, School 6)*

> *“I think social media has such a negative impact on what they deem as acceptable, you could literally go on Instagram now and type in sport, and there’d be girls walking around in next to nothing, sports bras and shorts.” (Female PE Staff, School 5)*

Pupils expressed that when their PE uniform was uncomfortable and ill-fitting, this could restrict their movement, reducing their level of participation in PE lessons. Additionally, some pupils described feeling either too cold (in winter) or too hot (in summer) in their uniform, which could also reduce level of exertion and participation. Some PE staff described adapting the PE uniform policy in these circumstances to try to enhance pupil comfort.

> *“The shorts are like ridiculously thick. Like I can barely breathe in these.” (Pupil, School 2)*

> *“the shorts sometimes like go up and it like distracts you into making sure your shorts don’t go up all the time. Because we have to like pull them down if they’re like getting really short.” (Pupil, School 6)*

> *“…in winter if we’re outside and it’s really cold they can wear a coat. They can bring gloves, they can wear a hat if they want to. I just want them to be comfortable. I want them to take part and that helps, definitely helps.” (Male PE staff, School 1)*

Although most PE staff appeared to value pupil choice and comfort, some also highlighted the importance of uniformity, with varying approaches to achieving this for PE uniforms across schools. Some schools offered unisex items (e.g. tops, shorts) to all pupils, to promote smartness and cater for diverse gender identities (see links to Theme 2). Other schools mandated school-branded items or plain, non-branded items to reduce social comparison between pupils.

> *“They were just unisex shorts and I think it’s all there and it’s easier kit wise and actually when they go to fixtures and different things they look quite smart. It’s not a scruffy kind of PE kit” (Female PE staff, School 6)*

> *“…more students maybe in terms of gender and we just want to make everyone feel comfortable that they didn’t have to pick one or the other.” (Female PE staff, School 2)*

> *“the uniform is in place so that no one feels uncomfortable in terms of brands they’ve got and stuff, whereas if we’re saying you can bring any jacket in, it’s going to be a brand war of what they’re allowed.” (Female PE staff, School 4)*

### Theme 2: PE uniforms are “made for boys”

Schools in our sample varied in whether they provided a universal, unisex or gender-based PE uniform. It appeared that many of the schools that provided a universal, unisex PE uniform (e.g. top and shorts) did this to ensure it fitted pupils of all gender identities, which was valued by some pupils.

> *“you just have it all that it fits boys, it fits girls and if you’re non-binary you can choose what you wear and it’s not like, ‘Oh you’re wearing a boys PE kit.’” (Pupil, School 3)*

> *“there’s two different types of joggers which are both unisex, the shorts are unisex, the t-shirts are unisex. So there’s no boys’ kit, girls’ kit, it’s just, ‘There’s our kit, pick and choose what you want to wear.’ Again, you’ve got so many kids at the moment that are gender fluid, or transitioning, or doing what they want, so you don’t want to be like, ‘This is the girls, that’s the boys.’” (Female PE staff, School 5)*

However, many girls and some PE staff expressed that unisex clothing often did not suit or fit female body shapes because it is *“made for boys”,* with unisex items being described as *“baggy”, “elongated”,* and *“not very flattering”*. When given choice, girls often opted for their own bottoms over the unisex options advertised on the school website. When unisex bottoms were compulsory, girls described needing to *“roll”* the shorts for a better fit (i.e. adjust the waistband). Some girls expressed that they disliked *“matching with the boys” (Pupil, School 1)*, and wanted *“some options for girls” (Pupil, School 6)*.

> *“they’re not very flattering at all and they don’t fit me very well or they don’t fit a lot of the girls that I know, very well. I feel like the shorts are made more for the boys than they are for the girls.” (Pupil, School 2)*

> *“lots of the girls won’t wear the shorts that we’ve got, because I don’t think they’re very flattering.” (Female PE staff, School 5)*

> ***“**Everyone just rolls them anyway… I wear shorts underneath but say you didn’t wear shorts underneath, if you don’t roll them, I feel like if you sit down you’re just going to flash everyone.” (Pupils, School 6)*

In schools with gender-based PE uniforms, items were typically advertised on uniform websites separately for boys and girls, despite some items being unisex. PE staff often described that their policy allowed pupils to choose any items. For example, the female PE staff in School 2 expressed *“if anyone wants to wear one or other kits, we’re absolutely fine with that”.* In contrast, girls often felt that the PE uniform rules meant that they were *“not allowed”* to wear items labelled for boys. Girls appeared to associate certain “boys” items (e.g. rugby tops) with particular sports, which they did not participate in and so felt they couldn’t wear. Despite some girls feeling that “boys” items would be more practical, they described feeling self-conscious wearing something labelled for boys because it would be against the norm (see links to Theme 1 and 3). For example, a pupil in School 2 expressed *“I don’t think some girls would be comfortable wearing the boys uniform… It’s just the name, knowing it’s boys uniform.”* Instead of unisex or gender-based uniforms, it was suggested by pupils that *“people would like having options on what fit to wear” (Pupil, School 1)*.

> *“**Participant:** there’s only joggers for boys. There’s none for the girls… **Interviewer:** You can’t wear joggers? **Participant:** We can but it’s only on the boys’ website. So if you go onto the uniform website there’s a boys’ section and girls’ section. In the boys’ section there’s joggers. In the girls’ section there’s just like skort and leggings.” (Pupils, School 3)*

> *“And they mostly do rugby all the time and their rugby t-shirts are normal gym tops… We don’t do rugby but the boys do rugby.” (Pupils, School 1) “certainly the girls don’t want to wear the rugby top at all” (Male PE staff, School 1)*

### Theme 3: Self-confidence influences comfort in wearing PE uniform, and in turn PE engagement

PE staff suggested that low self-confidence (i.e. lack of belief in oneself and one’s abilities) and negative body image were common issues among pupils, which became more noticeable with age. The Year 8 girls articulated varying levels of self-confidence and psychological comfort in their PE uniforms. Some girls expressed feeling self-conscious in their PE uniform, which led them to worry about how their appearance and ability would be perceived by others. In contrast, other girls were unconcerned with how they looked, allowing them to enjoy PE.

> *“you see other people and they look better in their P.E. kit than you do… even if you’re like good at sports and you enjoy sports and stuff, it’s just the P.E. kit makes you feel like they can do it better than you or something.” (Pupil, School 6)*

> *“I’ve never really thought about how I looked in the PE kit ‘cause I’ve always thought about the actual PE… I personally like the skort and t-shirt and as long as I’m okay with it I just don’t really care if other people look or if other people feel badly about it because I’m the one that’s wearing it.” (Pupil, School 2)*

> *“I just feel fine in my PE kit and it’s really comfy and I don’t feel like anybody’s looking at me.” (Pupil, School 4)*

Psychological comfort in wearing the PE uniform appeared to be influenced by body image concerns, judgement from others, and communal changing rooms, as illustrated by the quotes presented in Table 3. Some girls felt self-conscious in PE lessons due to concerns about their body hair and body size, leading them to want to cover up. It was suggested by pupils that many girls would avoid maximum exertion during PE because the PE uniform was uncomfortable when sweaty, but PE staff suggested this could be because getting sweaty would *“spoil”* their appearance (e.g. make-up and hair). PE staff suggested girls with lower sporting abilities were more appearance-conscious and less likely to engage in PE, unlike girls who enjoyed PE and felt confident in their ability. Fear of judgement about their appearance appeared to be a key concern for some girls, often heightened by negative comments, particularly from boys, which led some PE staff to keep boys and girls separate during PE. Self-consciousness appeared to be particularly exacerbated in communal changing rooms for some girls, with PE staff discretely allowing these pupils to change in other private places. In schools that allowed pupils to stay in their PE uniform all day, PE staff felt the policy had made a positive impact by eliminating the discomfort of changing in front of others.

**Table 3.** Factors influencing girls’ psychological comfort in wearing PE uniform and associated quotes linked to Theme 3 (Self-confidence influences comfort wearing PE uniform, and in turn PE engagement)

| **Factors influencing psychological comfort in PE uniform** | **Associated quotes** |
| --- | --- |
| Body image concerns, including body hair and body size | *"in the summer I'm just like really insecure about my body, like my body hair. So in the summer when it's boiling hot I don't really wanna wear a really short skirt because I'm not really secure with that." (Pupil, School 3)*<br>*"if you don't think you look good it's not really good for your self-confidence... for me like my shorts are really tight and everyone else's is really baggy so like it just makes me feel like, oh my thighs are thicker than hers and that like doesn't make me feel very good about myself." (Pupil, School 6)* |
| Sweatiness, appearance, and sporting ability | *"I feel if you have the collar it makes you sweat more as well and then people feel more uncomfortable in their PE kits and then they're less active to do PE." (Pupil, School 1)*<br>*"lower sets girls who are less able within PE, are more conscious of their appearance as well, and trying to get them to do PE is very difficult. Trying to get them to push themselves, to try and get them to get out of breath, because they don't want to spoil their makeup, they don't want to get sweat in their hair." (Female PE Staff, School 4)*<br>*"I feel confident in a lot of things I wear, I don't really mind what I look like and then because I enjoy P.E. as well so as long as I enjoy it I don't really mind what I look like in it." (Pupil, School 6)* |
| Fear of judgement | *“I feel like the PE kit itself is like, okay and I think a lot of people would be confident and wouldn't really care if it wasn't for the comments that other people make... if people are making comments and rude stuff like, I think it just lowers your confidence and you're just not gonna like it [the PE uniform] yourself.” (Pupil, School 4)*<br>*“we have to think about where the boys groups and girls groups are going to be because a lot of the girls are reluctant to sprint as hard as they can down the straight if the boys are near.” (PE staff, School 1)* |
| Communal changing rooms | *“I don't like changing clothes and like the changing room. I find that uncomfortable.” (Pupil, School 6)*<br>*“They do not like changing in the changing rooms in front of other people, which is fine if they don't. They can go to the toilets.” (Female PE staff, School 2)*<br>*“For me it's a much better system [referring to the all-day PE uniform policy] for many reasons. It's far more inclusive. You don't have issues with changing rooms and students who maybe don't feel so comfortable.” (Male PE staff, School 3)* |

### Co-developing PE uniform policies in secondary schools and ideas for recommendations

PE staff agreed that evidence-based, national-level guidance on school PE uniform policies would be valuable, providing guidelines are broad and adaptable, to account for varying school contexts and approaches.

> *“I think if there was a generic policy that supported schools who might need some guidelines, then it’s gonna be a benefit isn’t it” (Female PE staff, School 4)*

Pupils and PE staff across the six schools discussed ways to improve PE uniforms, which we have developed into initial recommendations for secondary schools to consider when reviewing and developing their PE uniform policies. Table 4 presents these initial recommendations by uniform item (tops, bottoms, additional layers, sports bras, and trainers/footwear), considering options, colour, style and fit, length, branding, and material, as well as, sizing, spare uniform, and affordability. Supplementary Table 3 presents these recommendations alongside supporting qualitative evidence.

**Table 4.** Initial recommendations for secondary school PE uniform policies.

| PE uniform policy category | Initial recommendations |
| --- | --- |
| Top | • The PE top should be the only mandatory PE uniform item and include a small school logo.<br>• Choose a neutral colour that is durable and can accommodate change in weather and temperature. Schools may wish to incorporate the school colour(s).<br>• Choose a well-fitting, round neck t-shirt without a collar.<br>• Choose durable, breathable, activewear material, avoiding mesh fabric (closely spaced holes) and too many seams.<br>• Choose a top that is comfortable and practical for physical activity rather than formal.<br>• Provide fitted and baggy options, alongside a regular unisex top for pupils to choose from, ensuring options are not labelled based on gender. |
| Bottoms | • School-branded bottoms should not be mandatory.<br>• Provide pupils with a choice, including for example, shorts, leggings, and tracksuit bottoms/joggers. Preferably allow pupils to wear their own bottoms. This will enable optimal comfort, style and fit.<br>• Consider providing rules or recommendations to pupils and parents about appropriate branding. Schools may wish to not allow branded items (e.g. Nike).<br>• Remove the option of skorts. Consider offering cycling shorts, but ensuring rules on appropriate length is mandated.<br>• Allow plain black bottoms, which is the easiest colour to find.<br>• Consider providing rules or recommendations to pupils and parents about appropriate activewear material for physical activity. |
| Additional layers | • Allow all pupils the option to wear underlayers/skins to cover their skin. Schools may wish to provide rules or guidance on appropriate colours, and branding. |
|  | • Consider allowing pupils to wear their own jackets and coats. Schools may wish to provide rules or guidance on appropriate colours, styles, and branding.<br>• Consider offering optional school branded hoodies or for pupils to wear their own hoodies for outdoor activities.<br>• Allow pupils to wear additional layers and religious clothing for religious reasons. Schools may also wish to provide rules or guidance on appropriate colours. |
| Sports bras | • Wearing a sports bra should be personal choice. Consider providing guidance on the importance of, and when pupils may need to start wearing, sports bras. |
| Trainers and footwear | • Provide guidance on appropriate styles and colours.<br>• Consider limiting the need for additional mandatory footwear, which is not regularly required during PE lessons (e.g. football boots) to reduce cost to parents. |
| Sizing | • Offer a large range of sizes for school branded items, and consider offering various styles (e.g. regular, fitted, baggy) to enable optimal comfort. |
| Spare uniform | • If possible, offer spare uniform that looks similar to the PE uniform, and ensure it is washed regularly. |
| Cost and affordability for parents | • Limit mandatory school branded items to the PE top only.<br>• Make it clear to pupils and parents from Year 7 which items are mandatory and must be school-branded, and which items pupils have the choice to wear their own clothing.<br>• Consider limiting branding allowed on own clothing to reduce cost, and pressure to buy 'trendy' items.<br>• Choose durable materials so uniform will not need to be replaced regularly.<br>• Consider collecting second-hand uniform from leavers and offering pre-loved items at a reduced price to support low-income parents. |

As highlighted in Theme 3, discomfort in communal changing rooms appeared to be an issue for many girls. Some PE staff also noted that *“girls can take ages to get changed” (Female PE staff, School 5)*, which reduced active time during PE lessons (as changing both in and out of PE uniform must happen during PE lessons). Some schools allowed pupils to wear their PE uniform all day (on PE lesson days) or come into or leave school in their PE uniform, reducing the need to change. This approach appeared to alleviate these issues, and the benefits and considerations discussed for adopting this policy are summarised in Supplementary Table 4.

Pupils emphasised the importance of being involved in updating PE uniform policies, noting *“we’re the ones wearing it, not them so they don’t know how it feels” (Pupil, School 2).* The PE staff agreed, with some describing previous ways of involving pupils, such as using school-wide voting systems and surveys. However, some staff preferred to involve a smaller *“select group”*, as involving all pupils could *“open up possible floodgates of like ‘We want this. We want that’” (Male PE staff, School 3).* Additionally, PE staff discussed the involvement of other stakeholders, including parents, Senior Leadership Teams (SLT), uniform designers and distributors, and for some schools, governors. While some PE staff felt SLT were *“supportive”* of them making PE uniform changes, others felt they had limited influence in changing current policies. Supplementary Table 5 presents a summary of the data, outlining stakeholder involvement in the PE uniform development process across schools.

PE staff highlighted that uniform policy changes would require at least one academic year to coordinate with uniform distributors, and gain school management approval. Both pupils and PE staff suggested implementing changes at the start of the academic year, introducing new mandatory PE items for Year 7 pupils only to minimise costs for older pupils.

> *“it would be almost like a year long process… the new year 7s coming in would have the new kit and then it would be open for the rest of the year groups to purchase if they wanted it. Rather than saying everyone’s got to have the new kit now. So just gradually include it into the policy.” (Male PE staff, School 1)*

## DISCUSSION

This qualitative study with Year 8 pupils and PE staff highlighted how PE uniforms can influence body image, and PE engagement among adolescent girls, and explored how PE uniform policies could be changed and co-developed to improve comfort for girls.

Our findings highlighted that PE uniforms that fit poorly, are too short, restrictive, and not weather appropriate (e.g. short skorts, tight collars, thin jackets) can discourage PE participation. These findings align with questionnaire-based research showing dissatisfaction among 14-16-year-old girls with PE uniform length and warmth,^25^ with 20% reporting PE uniform as a barrier to PE participation.^24^ Although limited and mixed, there is some quantitative evidence suggesting activity-enabling uniforms may improve physical activity levels. Using an ecological study design, comparing school-aged children’s physical activity levels across 135 countries/regions, Ryan et al., (2024)^42^ found that while school uniforms had no overall association with activity levels globally, they were associated with greater female gender inequalities in high-income countries (including the UK). Experimental studies in Australia comparing active versus traditional school uniforms in primary schools, collecting device-measured physical activity, found mixed results. One study reported increased activity for girls during break time^43^ and the other found minimal effects.^44^ Further research is required to determine whether redesigning secondary school PE uniforms can improve girls’ PE engagement and activity levels.

Our findings suggested that PE engagement among girls can be negatively impacted by uniforms that do not promote psychological comfort. When uniforms fit poorly (e.g. unisex designs) or expose too much skin, this can heighten feelings of self-consciousness, and amplify body image insecurities, such as body hair, and shape, as can changing in communal changing rooms. This appears to particularly impact girls with low self-confidence, potentially reducing their motivation and participation in PE, which can in turn further exacerbate poor body image. In contrast, self-confident girls appear less affected by PE uniforms. Our findings align with previous qualitative research with female adolescent athletes, which identified poor body image and inappropriate sportswear as major barriers to participation.^45^ In addition, a survey of 12-18-year-olds in Australia showed that more active girls were 20% more likely to report higher body satisfaction and greater willingness to wear any uniform.^26^ In our sample, 66% of Year 8 girls reported high levels of enjoyment in PE. However, a lack of self-confidence may be a more significant issue among girls with lower PE enjoyment and among older girls, as evidence indicates that body dissatisfaction tends to be higher among older and less active adolescents.^10^ ^46^ Furthermore, previous research highlights the links between self-confidence and self-esteem, and body dissatisfaction. For example, one study found that young females with lower self-esteem experienced greater body dissatisfaction when wearing tight, revealing sportswear compared to comfier baggy sportswear.^47^ Designing PE uniforms that promote self-confidence, particularly for girls who struggle with it, could play a key role in ensuring PE is enjoyable and inclusive for all girls.

Evidence suggests that image-based social media (e.g. Instagram, Tik Tok) is associated with body dissatisfaction and low self-esteem among adolescent girls as it may encourage social comparison and unattainable female body ideals.^4^ ^48^ This can negatively impact mental well-being and potentially contribute to disordered eating.^49–52^ Our findings suggested that female activewear fashion trends may influence girls’ clothing choices when allowed to wear their own clothes for PE. Striking the right balance between offering choice to promote psychological comfort, whilst minimising the focus on appearance and social comparison may be important for fostering self-confidence and enhancing PE engagement among girls.

Although research on how adolescent girls access and perceive online physical activity content is limited, one qualitative study suggested that engaging with fitness content can lead to setting unattainable body goals and body dissatisfaction among young adults.^53^ In addition, an experimental study found that browsing for activewear online lowered self-esteem, reduced positive body image, and heightened appearance comparison tendencies among women.^54^

### Implications for policy and future research

Governments advocate for gender-inclusive school and sports uniforms;^29–33^ however they lack specific recommendations for secondary schools in England. Our data suggests that recommendations should include offering a range of appropriate options for bottoms (e.g. shorts, leggings, joggers), providing various fits and styles without gender labels, allowing additional layers, and permitting pupils to wear PE uniform throughout the day on PE lesson days. These recommendations align with studies from Canada and New Zealand involving young active females, which also highlight the importance of flexibility, choice, inclusive sizing, comfortable materials, and styles designed for the female body.^26^ ^45^

Future research is needed with pupils, parents, PE staff and senior school staff to further develop and refine these recommendations to ensure they are fit for purpose. The findings also highlight a need to develop a national-level process by which schools can assess their current PE uniform policies, access evidence-based guidelines for improving comfort and inclusivity, and implement necessary changes. Once this is developed, it would be important to evaluate the impact on pupils’ body satisfaction, self-confidence, and engagement in PE.

### Strengths and limitations

By incorporating perspectives from both female pupils and PE staff and using activities to facilitate discussions, this qualitative methodology enabled us to explore beyond surface-level preferences for PE uniforms and understand in more depth how PE uniforms may influence girls’ body image and PE engagement. However, challenges in recruiting secondary schools resulted in a less diverse sample than planned, particularly with limited representation from schools in more deprived areas and pupils from ethnic minority backgrounds. While our initial PE uniform policy recommendations were developed using the data, only perspectives from Year 8 girls and PE staff in six mixed-sex Academy schools were captured. Further research is required to ensure these recommendations are inclusive for all pupils, including those with different gender identities, Special Education Needs and Disabilities (SEND) and from ethnic minority backgrounds. Additionally, most girls in our sample reported high levels of PE enjoyment, highlighting the need to ensure future recommendations are suitable for girls with low PE engagement.

## CONCLUSION

This qualitative study highlights the importance of having secondary school PE uniform policies that promote comfort and self-confidence among adolescent girls to facilitate better PE engagement. Our findings suggest it is important to balance offering pupils choice over PE uniform items to facilitate better comfort, whilst also maintaining uniformity to reduce social comparison and ensure items are appropriate for pupils of different gender identities. Girls who lack self-confidence may be more self-conscious about how they look in their PE uniform, which may reduce PE engagement. As well as offering unisex items, schools should consider offering a variety of styles and sizes without gender labels to ensure uniforms fit female bodies, as well as allowing pupils to wear their own bottoms and additional layer to help improve self-confidence.

## Data Availability

Anonymised transcripts will be made available on the University of Bristol Data Repository (data.bris) under the restricted access data policy.

https://www.bristol.ac.uk/staff/researchers/data/accessing-research-data/

## Contributors

AP, LC, CC, RW, SB, IPV, AA, RJ, and HB were responsible for study conceptualisation. AP and EC were responsible for project administration. AP and EC were responsible for recruitment. AP, EC and LC were responsible for data collection. AP, EC, LC and CC were responsible for data analysis. AP wrote the first draft of the manuscript. All authors contributed to and approved the final manuscript.

## Funding

This work was funded by The National Institute for Health Research, Bristol Biomedical Research Centre, grant number NIHR203315. The views and opinions expressed are those of the authors and do not necessarily reflect those of NIHR or the Department of Health and Social Care. This work is independent research supported by the National Institute for Health and Care Research Yorkshire and Humber Applied Research Collaboration. The views expressed in this publication are those of the author(s) and not necessarily those of the National Institute for Health and Care Research or the Department of Health and Social Care.

## Acknowledgments

We would like to thank the participants for taking part in the study. We would also like to thank PPIE members for their input into study development.

## Conflicts of Interest

None

## Ethical Approval Statement

The Faculty of Health Sciences Ethics Committee at the University of Bristol granted ethical approval (Reference number: 17624).

## Data sharing

Anonymised transcripts will be made available on the University of Bristol Data Repository (Accessing data in data.bris | Staff | University of Bristol) under the restricted access data policy.

## Notes

### Competing Interest Statement

The authors have declared no competing interest.

### Clinical Protocols

https://osf.io/krhg7

### Author Declarations

Ethics approval was obtained from the Faculty of Health Sciences Ethics Committee at the University of Bristol, in February 2024 (Reference 17624)

### Summary of Updates

This version of the manuscript has been revised to correct the author list

