## Supplementary Table 5 for "A qualitative study exploring how secondary school PE uniform policies influence body image, and PE engagement among adolescent girls"

**Supplementary Table 5. How stakeholders can be involved in developing PE uniform policies with associated evidence**

| Who is involved | Summary of how they can be involved | Evidence from focus groups/interviews |
|---|---|---|
| Pupils | • It may be important for schools to consider changes to the PE uniform if pupils raise concerns about lack of comfort.<br><br>• Pupil opinions on changes to the PE uniform can be expressed using voting systems or surveys, or a select group of pupils (e.g. ambassadors) could be asked to gather opinions from wider groups of pupils. Opinions collected should be representative of the school as a whole.<br><br>• A representative group of pupils can try on samples of new PE uniform items, and provide feedback. | In some schools, changes to the PE uniform had been made following pupil requests, such as options to wear leggings and material of the PE tops<br><br>*"Things like sports leggings for the girls became something that was really popular and it was something that – we listened to them" (Male PE staff, school 1)*<br><br>Some PE staff described using a select representative group to discuss potential changes, as well sports ambassadors or sports captains, as it would be difficult to please everyone.<br><br>*"**Participant:** I'm going to have a look at different materials. We've had requests for different materials for the tee shirt…"… **Interviewer:** "How did you select the pupils to be involved in that?" **Participant:** "I had arranged different year groups, some team players, some non-team players, different sizes…" (Female PE staff, school 6)*<br><br>*"I wonder if we put something about kit it would open up possible floodgates of like, 'We want this. We want this. We wanna wear Nike.' So I think in that case you do a slightly more select group" (Male PE staff, school 3)*<br><br>Pupils suggested that a petition or vote would be effective in communicating their opinions, for example, in one of the schools, pupils had voted for their top colour.<br><br>One school used uniform samples to allow some pupils to try out possible new uniforms, and then used a majority vote to help consider what changes would be most liked by pupils and parents. |

| | | |
|---|---|---|
| Parents | • Schools can ask for feedback from parents on any potential changes to the PE uniform, especially if changes have cost implications. Any changes can be communicated to parents before the start of an academic year via school communication channels (e.g. online portals, letters home, newsletters, forums, and emails). | PE staff stated the importance of involving parents when making changes to the PE uniform, as parents can refuse to buy new uniform. PE staff suggested factors such as price, durability, and whether the uniform can be passed down or shared with younger siblings were most important to parents.<br><br>*"Yeah, I think inexpensive and something that is easy to wash, easy to clean, that type of thing would be something that the parents would be keen to do and I think you're right, I think getting parents voice onboard would be crucial to this " (Male PE staff, school 1)*<br><br>*"So then also some parents might want the younger ones to have the same kit that the older ones have had and just hand it down, so it does take a while to kind of filter through everything." (Female PE staff, school 6)*<br><br>Most staff described that they would communicate any changes being considered or made to the PE uniform via email or newsletters.<br><br>*"so they will obviously get information letters et cetera. It's all on the school website as well, the information. They'll get emails home for the whole year groups that this is happening. Yeah, communication is generally done through email with all the parents." (Female PE staff, school 2)*<br><br>The use of a parent forum was also mentioned, where parents could give feedback on proposed PE uniform changes. |
| PE staff | • It may be important for PE staff to discuss concerns about the PE uniform with pupils.<br><br>• PE staff could be given the autonomy and responsibility to propose changes to the PE uniform policy by the Senior Leadership Team (SLT), and | In some schools, the process of changing the PE uniform involved PE staff first discussing their ideas with the Head of department, which were then taken to SLT before being approved.<br><br>*"Our Head of PE is really good, if you go to her with something then she's very open, and she will take into account all different things. And if we said, 'We* |

| | | |
|---|---|---|
| | to liaise with uniform designers and distributors. | *want to do this,' she would go to the SLT, and she will fight our corner." (Female PE staff, school 5)*<br><br>In other schools, PE staff described SLT requesting changes to, and making decisions about the PE uniform that were not discussed with the PE department in advance, which they found frustrating.<br><br>*"If we wanted to, we probably have to [pause] give it to SLT and be like, 'This is what we want to do. Can we do it?' and they have the final say… Well, that kit changed, and we had no say in it whatsoever."* (Female PE staff, school 5) |
| Senior Leadership Team | • SLT and PE staff could discuss proposed changes to the PE uniform together before approval. | Some PE staff described having supportive SLT who would take on suggestions from the PE department. However, others expressed that they were not in support of the decisions made by SLT regarding the PE uniform, as they were not based on the needs of pupils or the PE department.<br><br>In some schools, the policy to allow pupils to wear their PE uniform all day was introduced during COVID-19. This policy was continued post-COVID in some schools, which PE staff were supportive of. However, other schools had reverted back to changing, which some PE staff felt they'd had little input into.<br><br>*"**Interviewer:** How easy was it to get the senior leadership on board with the idea to keep the [all-day PE uniform] policy? **Participant:** Pretty easy. I think they were very supportive of it; I think they saw that it worked well." (Female PE staff, school 4)*<br><br>*"So we're like, 'Oh this great, we can – but we tried to keep it. Like, 'Can we keep this?' But the Senior Leadership Team were, 'No, this is not happening.'" (Female PE staff, school 2)* |

| | | |
|---|---|---|
| Uniform designers and distributors | • Schools must liaise with uniform designers and distributors when making changes to the PE uniform, as they may have limited options for designs, styles, materials, sizing etc., and may want old stock to be sold before introducing new designs, which could delay changes.<br>• Uniform designers and distributors may need to consider the gender labelling of their clothing. | PE staff described that they would work with their uniform designers and distributors to make any changes, however this sometimes limited the options for design, fit, and material.<br><br>Pupils and staff both highlighted that if changes are made to the PE uniforms, uniforms should be introduced gradually, and made mandatory only for new Year 7 pupils, so that buying leftover stock was still an option, reducing waste, and so there is less pressure on parents to buy new PE uniform<br>*"we were too late for September but I think if we get moving before Christmas then I think we could easily be able to change it for the following September, but I think it would be almost like a year long process of working out, A, what we want, looking at different companies, what they can offer and then going from there*" *(Male PE staff, school 1)*<br>*"introducing it from the new cohort that comes in, with the option of giving it to the older students as well if they wanted to." (Female PE staff, school 4)* |
| Governors* | • In some schools, changes to the PE uniform policy will require approval from Governors, which needs to be considered in the timeline of implementing changes. | One school mentioned that Governors must be involved in the process of making "big" changes to school policies *"–that would be the first step; SLT then the Head, then the governors." (Female PE staff, school 6)* |

*Governors are a selected group of individuals accountable for a school, operating at a strategic level
