## Supplementary Table 4 for "A qualitative study exploring how secondary school PE uniform policies influence body image, and PE engagement among adolescent girls"

**Supplementary Table 4. Benefits and considerations for schools adopting a PE uniform policy to eliminate or reduce the need to change for PE**

| **Allow pupils to wear PE uniform all day, on days they have PE** | | **Allow pupils to come into school or leave school in their PE uniform, on days they have PE in the morning or afternoon** | |
|---|---|---|---|
| **Pros** | **Cons** | **Pros** | **Cons** |
| Removes feelings of discomfort, insecurity and self-consciousness when changing in front of peers in communal changing rooms, which are often described as unpleasant places | Pupils are limited to doing PE indoors when it is raining, as they may not have a change of clothing | Reduces feelings of discomfort, insecurity and self-consciousness when changing in front of peers in communal changing rooms, which are often described as unpleasant places to only once per PE class | Some pupils may try to bend the rules by wearing their PE uniform on non-PE days, which staff must monitor and sanction |
| Promotes inclusivity for pupils with diverse gender identities who might otherwise be expected to choose between binary changing room options | PE uniform can smell if worn all day after doing PE or if not washed on a regular enough basis | Increases time for activities during PE and breaks | Some pupils may forget or purposely not bring their school uniform to change into after PE, which staff must monitor and sanction |
| Increases time for activities during PE and breaks | Some pupils may try to bend the rules by wearing their PE uniform on non-PE days, which staff must monitor and sanction | Pupils are less likely to forget their PE uniform if worn into school, which reduces PE staff time spent sourcing spare uniform, and the need to sanction forgotten PE uniform | May be inconvenient to carry school uniform in a bag to change into after PE |
| More convenient for pupils, as they don’t need to carry their PE uniform to and around the school | Uniform may look less formal and consistent across the school | Pupils are less likely to be late to catch the bus home | Uniform may look less formal and consistent across the school |

| | |
|---|---|
| Pupils are less likely to forget their PE uniform if worn into school, which reduces PE staff time spent sourcing spare uniform, and the need to sanction forgotten PE uniform | Pupils who feel uncomfortable in their PE uniform may not have the option to change |
| Pupils are less likely to be late to lessons or miss the bus home | |
| PE uniform is often comfier than the regular school uniform | |
| Belongings such as phones, jewellery, and items of clothing are less likely to go missing | |
| Pupils are more comfortable wearing sports bras as they don’t have to change in and out of them in front of others | |
