## Supplementary Table 3 for "A qualitative study exploring how secondary school PE uniform policies influence body image, and PE engagement among adolescent girls"

**Supplementary Table 3. Initial recommendations for secondary school PE uniform policies with associated qualitative evidence**

| | **Recommendations** | **Evidence from qualitative focus groups and interviews** |
|---|---|---|
| **Tops** | | |
| Branding | • The PE top should be the only mandatory PE uniform item and include a small school logo. | Pupils and staff agreed that the PE top should be worn to represent their school.<br>*"When it's on there, it proves that you're from that school and it shows – in like sports days and that and it's really good."* (Pupil, School 2) |
| Colour | • Choose a neutral colour that is durable and can accommodate change in weather and temperature. Schools may wish to incorporate the school colour(s). | Dark colours such as black and navy were often described as too hot, especially in the summer, as the colour absorbs heat. A white top was described as cooler due to reflecting the light, however it could be see-through and less durable (e.g. dirt shows up, fabric changes colour after washing).<br>*"it goes like grey. 'Cause you gotta wash it really regularly, it just goes a horrible colour."* (Pupil, School 1)<br>Pupils and staff wanted the PE top to represent the school, and generally liked the idea of having school or house colours represented in the PE tops (e.g. colour stripes representing their school or house colour). However, it may not be possible to pass down tops with house colours to younger siblings.<br>*"in year 7, whatever house they're in that's the colour stripe they buy and then they don't have to put bibs on et cetera. But the parents were very much like, 'No.' They didn't like it for reasons, if you've got a younger child or if it's swapping a house for example."* (Female PE staff, School 2) |

| | | |
|---|---|---|
| Collar | • Choose a well-fitting, round neck t-shirt without a collar. | Most pupils in schools with button-up collared polo shirts disliked their PE top, as they described it to be *"itchy", "tight", "uncomfortable"*, and *"sweaty".*<br><br>Some pupils felt uncomfortable that the neck of their tops were too baggy, and felt too low down on their chests.<br><br>*"the bra can show 'cause of how like low it is." (Ppupil, School 4)* |
| Material | • Choose durable, breathable, activewear material, avoiding mesh fabric (closely spaced holes) and too many seams. | Pupils in some schools disliked cotton PE tops because the material was too *"thick", "hot", "sweaty"*, and *"not easy to wash".*<br><br>A few pupils mentioned that mesh fabric (closely spaced holes) can be *"uncomfortable", "itchy"*, and cause skin irritation, as well as *"loads of seams"* causing *"a sensory issue".* |
| Style and fit | • Choose a top that is comfortable and practical for physical activity rather than formal.<br><br>• Provide fitted and baggy options, alongside a regular unisex top for pupils to choose from, ensuring options are not labelled based on gender. | Pupils and staff felt that it was more important for PE uniform to be practical and comfortable, than formal.<br><br>Pupils wanted choice over the fit of the top, as some preferred fitted and some preferred baggy. They expressed that fitted tops should not be advertised for girls only, and baggy tops for boys only, to allow everyone choice over which fit they prefer, without the fear of standing out.<br><br>*"I think people would like having options on what fit to wear." (Pupil, School 1)*<br><br>*"It's like it's not fitted but it's not baggy. Like it's really annoying 'cause I don't want it to be like fitted so it's like carving your like body but…" (Pupil, School 4)* |
| **Bottoms** | | |

| | | |
|---|---|---|
| Options (for style and fit) | • Provide pupils with a choice, including for example, shorts, leggings, and tracksuit bottoms/joggers. Preferably allow pupils to wear their own bottoms. This will enable optimal comfort, style and fit. | Many pupils felt the school branded options for bottoms were *"ugly",* unflattering, *"too baggy", "too tight"*, or did not suit their shape. Pupils often described waistbands being too big, and shorts being too baggy, especially if bottoms were unisex. For example, they described needing to *"roll"* or adjust them to fit, suggesting that it would be difficult to provide school branded options for bottoms that would suit everyone.<br><br>*"Everyone just rolls them anyway. I roll them about three times and they're so long"* (Pupil, School 6)<br><br>Pupils and staff felt offering choice over bottoms helped improve comfort and inclusivity. Pupils liked to choose bottoms they felt most comfortable wearing, which was sometimes weather and activity dependent.<br><br>*"I like that you can wear leggings and shorts depending on the weather"* (Pupil, School 1) |
| Branding | • School-branded bottoms should not be mandatory.<br>• Consider providing rules or recommendations to pupils and parents about appropriate branding. Schools may wish to not allow branded items (e.g. Nike). | Limiting school branded PE uniform items reduces the cost to families. If choice of own clothes is given alongside school branded options, pupils will often opt to wear their own clothes, which can mean parents spend money on buying the full school branded PE uniform for their children in Year 7, which is no longer worn in Year 8 onwards.<br><br>*"in year 7 I hated it. I hated PE because I hated the uniform, but now I'm like more okay with it and I care less because I started wearing more of leggings and making more my own choices"* (Pupil, School 2)<br><br>Some pupils and staff felt limiting fashion branding helped to promote equality so that pupils would not compare or judge each other, however, others felt that branding did not cause issues. |

| | | |
|---|---|---|
| | | *"I think they don't allow us to wear leggings because if you've got like I guess expensive leggings… And then somebody doesn't have stuff like that, it's going to make you feel like you're not equal to them, that's why I think everyone's got similar."* (Pupil, School 6) |
| Length | • Remove the option of skorts. Consider offering cycling shorts, but ensuring rules on appropriate length is mandated. | Most pupils who wore or could wear skorts as part of their PE uniform felt uncomfortable about how short they were and felt skorts restricted the activities they could do in PE. In addition, a few pupils felt being allowed to wear a short skort contradicted school rules about length of skirt allowed for their regular school uniform.<br><br>*"The skorts are really short… When I'm wearing I'm like, 'Why is this not longer.'"* (Pupils, School 3)<br><br>Cycle shorts were very popular among pupils, and most wanted their school to allow them, however staff had concerns about the appropriateness and length of cycle shorts.<br><br>*"It's not practical for all activities. Some of them would go ridiculously short and that's the last thing you want to see." (Female PE staff, School 6)* |
| Colour | • Allow plain black bottoms, which is the easiest colour to find. | Most schools in the study had the choice of black bottoms. Pupils and staff felt black was practical and easy to purchase. Pupils expressed feeling comfortable wearing the colour black.<br><br>*"I think black's quite a good colour 'cause it suits everyone. So a lot of people feel comfortable in it."* (Pupil, School 1)<br><br>In a school where navy bottoms were part of the PE uniform, pupils said it was sometimes difficult to find in shops, and tended to be more expensive than black bottoms. |

| | | |
|---|---|---|
| | | *"sometimes navy is a bit of a harder colour to find in joggers. And I think black is such a similar colour to navy that I think you should just be allowed black ones."* (Pupil, School 4) |
| Material | • Consider providing rules or recommendations to pupils and parents about appropriate activewear material for physical activity. | Pupils disliked bottoms made from thick material, as this becomes too hot, especially in the summer, and is difficult to wash and dry.<br><br>Staff suggested that activewear material was most appropriate to ensure bottoms are not see-through.<br><br>*"It can't be something you've got from Primark, or leggings from Asda, it has to be the proper material. Because they do wear the cheaper ones and they become see-through, and when they're moving around"* (Female PE staff, School 5) |
| **Additional layers** | | |
| Underlayers/skins | • Allow all pupils the option to wear underlayers/skins to cover their skin. Schools may wish to provide rules or guidance on appropriate colours, and branding. | Schools allowed pupils to wear additional layers for religious reasons but varied in their policies about whether underlayers/skins was allowed for all pupils.<br><br>Pupils expressed that they would like the option to wear additional layers to keep them warm. In addition, some pupils felt self-conscious about showing their skin and bodies and wanted a way to cover up, especially when participating in certain activities that might expose their skin (e.g. trampolining, gymnastics).<br><br>*"we do wear like long sleeve like white tops and then that's people who don't like their arms but they would have to ask to wear them."* (Pupil, School 6) |
| Hoodies/jackets/coats | • Consider allowing pupils to wear their own jackets and coats. Schools | Some pupils complained about not being allowed to wear coats, jackets or jumpers when doing PE outside in the cold. Pupils wanted to be able |

| | | |
|---|---|---|
| | may wish to provide rules or guidance on appropriate colours, styles, and branding.<br>• Consider offering optional school branded hoodies or for pupils to wear their own hoodies for outdoor activities. | to wear their own coats during PE, however some staff felt certain coats were not practical for physical activity (e.g. puffa jackets).<br>Pupils often felt school branded jackets were not fit for purpose because they were too baggy or too thin, and therefore did not keep them warm. Some pupils disliked zip-up jackets with collars because they were unflattering and uncomfortable.<br>*"the coat is way over priced considering it is as thin as a sheet of paper" (Pupil, School 4)*<br>Pupils wanted to be able to wear hoodies, which they felt were most comfortable and warm, but also because they were fashionable.<br>*"why can't they just make a hoodie. I get kind of get why not having your hoods on in class but during break and stuff, it's just nicer. These are just small, hoodies are just baggier and nice." (Pupil, School 3)* |
| Religious clothing | • Allow pupils to wear additional layers and religious clothing for religious reasons. Schools may also wish to provide rules or guidance on appropriate colours. | All schools allowed pupils to wear religious clothing and additional layers for religious reasons during PE lessons.<br>*"It's an adaptable thing… they will wear maybe a traditional sort of dress and they'll take part in PE. It will be quite baggy but they can still take part and we wouldn't ever sanction that." (Male PE Staff, School 3)* |
| **Sports bras** | • Wearing a sports bra should be personal choice. Consider providing guidance on the importance of, and when pupils may need to start wearing, sports bras. | Pupils and staff agreed that pupils should choose whether to wear a sports bra or not but that guidance would be helpful.<br>*"To be fair, no we don't talk to them about sports bras. That's a really good thing actually, I've never even thought about it. There are girls that* |

| | | |
|---|---|---|
| | | *play sport, do PE, and we're expecting them to run around and they're probably just wearing a normal bra." (Female PE staff, School 4)*<br><br>Some pupils expressed discomfort about wearing sports bras for PE because of having to change in and out of them in front of peers.<br><br>*"Some people don't wanna wear it for the entire day. You don't wanna get to the time where it's PE and then have to change your bra"* (Pupil, *School 2)* |
| **Trainers and footwear** | • Provide guidance on appropriate styles and colours.<br>• Consider limiting the need for additional mandatory footwear, which is not regularly required during PE lessons (e.g. football boots) to reduce cost to parents. | Pupils liked that they could wear their own trainers. Staff expressed it was important to provide guidance on inappropriate footwear, such as not allowing fashion trainers, as these are not practical for physical activity.<br><br>*"we don't allow like Jordan 1s and stuff like that, that's like purely fashion trainers that actually aren't good for sport." (Male PE staff, School 3)* |
| **Sizing** | • Offer a large range of sizes for school branded items, and consider offering various styles (e.g. regular, fitted, baggy) to enable optimal comfort. | Some schools had a large range of sizes available to suit everyone, whilst some pupils and staff in other schools felt that larger size ranges were not available.<br><br>Some pupils felt that unisex PE uniform items were ill-fitting and did not suit all body types.<br><br>*"My friend in year 7, she tried to get the skort or the shorts but she's quite a big build and she couldn't find a size that fits her, so now she just* |

| | | |
|---|---|---|
| | | *has to wear cycling shorts and she gets a detention every time." (Pupil, School 3)* |
| **Spare uniform** | • If possible, offer spare uniform that looks similar to the PE uniform, and ensure it is washed regularly. | Some pupils expressed feeling self-conscious when having to wear spare uniform because it made them look different to everyone else.<br>*"it can affect your lesson 'cause you don't feel confident in what you're wearing and then you'll put yourself down the whole lesson 'cause you'll keep blaming yourself that you forgot your PE kit and you're the only one that's stands out in front of everybody else." (Pupil, School 1)*<br>Some PE staff described collecting pre-loved uniform items and keeping these washed and available for pupils who forgot their uniform, which helped with PE engagement.<br>*"we do have spare kit, so I'm constantly doing washing but we've got quite a collection now of like shorts and t-shirts and all our old trainers and different things or trainers that others have grown out of. So most of them are quite good and go "Oh miss I'm really sorry, please can I borrow a t-shirt?"" (PE staff, School 6)* |
| **Cost and affordability** | • Limit mandatory school branded items to the PE top only.<br>• Make it clear to pupils and parents from Year 7 which items are mandatory and must be school-branded, and which items pupils | Pupils and staff felt it was important that PE uniform was not too expensive. It was expressed that allowing pupils to wear their own bottoms and additional layers could reduce cost to parents. Some pupils expressed it was unnecessary for their parents to buy the non-mandatory PE uniform items (e.g. bottoms) in Year 7 because if they had the choice, they would opt to wear their own clothing. |

| | | |
|---|---|---|
| | have the choice to wear their own clothing.<br>• Consider limiting branding allowed on own clothing to reduce cost, and pressure to buy ‘trendy’ items.<br>• Choose durable materials so uniform will not need to be replaced regularly.<br>• Consider collecting second-hand uniform from leavers and offering pre-loved items at a reduced price to support low-income parents. | *“a lot of people would already have just a plain black jumper or summut like that. So it’s more accessible because it doesn’t mean that people have to buy more… ‘Cause some people might not be able to afford it.” (Pupils, School 1)*<br>Some staff described strategies they used to reduce the cost of the PE uniforms for families of low income, including selling pre-loved items at discounted prices, giving certain pupils spare unclaimed uniform, and using Pupil Premium budget to subside cost of uniform to certain parents.<br>*“we ask the year 11s when they leave for any kits… So they’d sell it for like £1.50 a top and things like that and stuff. So just try and make it so everyone can afford it. But there’s not always lots. I think we’ve ran out already. Normally in the transition days we’ll have lots of kit that is available to buy and things.” (PE staff, School 2)* |
