## Supplementary Table 2 for "A qualitative study exploring how secondary school PE uniform policies influence body image, and PE engagement among adolescent girls"

**Supplementary Table 2. Summary of PE uniform policies by school**

| School | PE uniform policy accessible | Discrepancies between PE uniform policy and focus groups/interviews | Unisex uniform | Options for girls (e.g. leggings) | Top colour | School had all-day PE uniform policy |
|---|---|---|---|---|---|---|
| 1 | Stated on school website | Pupils can wear branded items despite policy stating that this is not permitted<br>Pupils can wear leggings despite policy not permitting | Yes | Allowed to wear own leggings, with no strict policy on branding | White | No |
| 2 | Stated on school website | Policy states that pupils are not permitted to wear cycle shorts, however this is sometimes allowed | "No gender restrictions" | Allowed to wear own plain black sports shorts or leggings, with no branding<br>School skort was an option | White or black | No |
| 3 | Stated on school website | None | Yes | Allowed to wear plain black leggings or school branded leggings, skort and tracksuit bottoms | Black | Yes |
| 4 | Stated on school website | None | Choice between unisex or female fit for tops and shorts | Allowed to wear plain navy sports leggings or school branded leggings or tracksuit bottoms | Navy and blue | Yes |
| 5 | Stated on school website | None | Yes | Allowed to wear plain black or navy leggings with discrete branding or school branded unisex navy joggers or tracksuit bottoms | Blue | No |
| 6 | No – access only via school portal | N/A | Yes | Not allowed to wear own clothing. School-branded shorts or tracksuit bottoms given as options | White | No |
